# COVID-19 COGNITIVE DEFICITS AFTER RESPIRATORY ASSISTANCE IN THE SUBACUTE PHASE: A COVID-REHABILITATION UNIT EXPERIENCE

**DOI:** 10.1101/2020.11.12.20229823

**Authors:** Federica Alemanno, Elise Houdayer, Anna Parma, Alfio Spina, Alessandra Del Forno, Alessandra Scatolini, Sara Angelone, Luigia Brugliera, Andrea Tettamanti, Luigi Beretta, Sandro Iannaccone

**Author notes:** Correspondence: Dr. Elise Houdayer, Department of Rehabilitation and Functional Recovery, IRCCS San Raffaele Hospital, via Olgettina 60, 20132, MI, Milan, Italy.

## Abstract

**Introduction:** COVID-19 complications can include neurological, psychiatric, psychological, and psychosocial impairments. Little is known on the consequences of SARS-COV-2 on cognitive functions of patients in the sub-acute phase of the disease. We aimed to investigate the impact of COVID-19 on cognitive functions of patients admitted to the COVID-19 Rehabilitation Unit of the San Raffaele Hospital (Milan, Italy).

**Material and Methods:** 87 patients admitted to the COVID-19 Rehabilitation Unit from March 27^th^ to June 20^th^ 2020 were included. Patients underwent Mini Mental State Evaluation (MMSE), Montreal Cognitive Assessment (MoCA), Hamilton Rating Scale for Depression, and Functional Independence Measure (FIM). Data were divided in 4 groups according to the respiratory assistance in the acute phase: Group1 (orotracheal intubation), Group2 (non-invasive ventilation using Biphasic Positive Airway Pressure), Group3 (Venturi Masks), Group4 (no oxygen therapy). Follow-ups were performed at one month after home-discharge.

**Results:** Out of the 87 patients (62 Male, mean age 67.23 ± 12.89 years), 80% had neuropsychological deficits (MoCA and MMSE) and 40% showed mild-to-moderate depression. Group1 had higher scores than Group3 for visuospatial/executive functions (p=0.016), naming (p=0.024), short- and long-term memory (p=0.010, p=0.005), abstraction (p=0.024), and orientation (p=0.034). Group1 was younger than Groups2 and 3. Cognitive impairments correlated with patients’ age. Only 18 patients presented with anosmia. Their data did not differ from the other patients. FIM (<100) did not differ between groups. Patients partly recovered at one-month follow-up and 43% showed signs of post-traumatic stress disorder.

**Conclusion:** Patients with severe functional impairments had important cognitive and emotional deficits which might have been influenced by the choice of ventilatory therapy, but mostly appeared to be related to aging, independently of FIM scores. These findings should be integrated for decision-making in respiratory and neuropsychiatric assistance of COVID-19 patients in the subacute phase of the disease, and show the need for long-term support and psychological treatment of post-COVID-19 patients.

## INTRODUCTION

The coronavirus disease 2019 (COVID-19) pandemic has dramatically shaken the healthcare system, worldwide. The severe acute respiratory syndrome coronavirus 2 (SARS-CoV-2) responsible for the COVID-19 can be responsible for various clinical features, ranging from asymptomatic to critical health conditions. Among these features, the most common forms are: (1) mild, with no dyspnea, no low blood oxygen saturation (SatO_2_); (2) moderate, with dyspnea, SatO_2_= 94% to 98%, radiological signs of pneumonia; (3) severe, with dyspnea, SatO_2_ ≤ 93%, respiratory rate (RR) > 30/min, radiological progression of lesions, with O2 supplementation required, eventually with non-invasive ventilation; and (4) critical with patients needing mechanical ventilation. [1] In all these various clinical conditions, patients can present with cardiorespiratory, neurologic or systemic complications, leading to the need for functional rehabilitation for about 20% of hospitalized COVID-19 patients. [1–4] Nonetheless, in the first months, hospitals tended to discharge patients as soon as possible to face the increasing needs for hospital admissions. A communication from the San Raffaele Hospital of Milan reported that, in the exponential phase of the national pandemic, about 25% of patients needed specialized rehabilitation to address cardiorespiratory, motor and/or cognitive dysfunctions in the subacute phase. [4] Indeed, in this subacute phase (from five to twenty days after symptoms onset), patients are still infectious for COVID-19 and can be in need for functional rehabilitation. The clinical care of these patients should be organized according to the clinical status and symptoms of the patients. Recommendations have been addressed to support the implementation of a multidisciplinary rehabilitative pathway for those COVID-19 patients in need for functional recovery. [1,4] According to this new organization, COVID-19 patients with functional deficits (with positive swab and no need for ICU) should be transferred to dedicated COVID-19 rehabilitation units when they meet the following criteria: stable for at least three days (no recurrence of fever, both respiratory rates (RR) and SatO2 stable, radiological progression of the disease has been ruled out) and Functional Independence Measure (FIM) shows areas of dependence (score < 100[5]). Neurological complications of the COVID-19, such as dizziness, headache, ageusia or anosmia, have been described. [6–8] Patients can also suffer from signs of deconditioning, critical-illness-related myopathy and neuropathy (CRIMYNE), dysphagia, joint stiffness, and pain. [1] It has also been recently shown that patients can present sub-clinical signs of neuronal suffering, even in the absence of neurological symptoms. [9] Psychiatric complications have also been reported, such as encephalitis, cerebrovascular disease (ischemic stroke or intracerebral hemorrhage), psychosis or neurocognitive syndrome (dementia-like). [6,10–13]

Reports have also been made on the psycho-social effects of the COVID-19 pandemic on patients, caregivers and on the general population in relation with home-confinement. Most of these communications reported issues related to anxiety, depression, and post-traumatic stress syndrome.[14,15]

Although neurological, psychiatric, and psychological signs have been reported in COVID-19 patients, there is a lack of data regarding the actual consequences of the disease on the cognitive functions in patients still presenting signs of SARS-CoV-2 infection. In this study, we aimed to investigate the impact of COVID-19 on the cognitive functions of infectious patients admitted to the COVID-19 Rehabilitation Unit of the San Raffaele Scientific Institute of Milan (Italy), in the sub-acute phase of the disease (about ten days after symptoms onset). For data analysis, patients were separated in four different groups according to the type of respiratory assistance they benefited in the acute phase of the disease.

## MATERIALS AND METHODS

### Population

Consecutive patients admitted to the COVID-19 Rehabilitation Unit of the San Raffaele Scientific Institute (Milan) from March 27^th^ to June 20^th^ 2020 have been included. Criteria to admit COVID-19 patients in this Unit were: positive swab for SARS-CoV-2, stable SatO_2_ and RR, no need for respiratory assistance or no more than two l/min, absence of fever, and with areas of dependence at the FIM evaluation (FIM score < 100).[4] These patients had been previously admitted in the Emergency Room (ER), Intensive Care Units (ICU), Respiratory High Dependency Care Units (RHDCU) or Infectious Diseases units of the San Raffaele Hospital. We excluded from the subsequent clinical study patients who were treated for cognitive dysfunctions, patients who were under psychotropic drugs prior to their recovery, and patients presenting with COVID-19 encephalitis. Patients with a disease onset less than five days and superior to 20 days were also excluded (patients were thus in the sub-acute phase of the disease, five to twenty days after symptoms onset). Oral and written consents were obtained from participants, in accordance with the Code of Ethics of the World Medical Association (Declaration of Helsinki) and the study was approved by our local Ethics committee.

### Evaluations and measurements

At their admission in the COVID-19 Rehabilitative Unit, patients underwent neuropsychological evaluation including: Mini Mental State Evaluation (MMSE)[16], Montreal Cognitive Assessment (MoCA)[17], and Hamilton Rating Scale for Depression (HRSD)[18]. No further detailed neuropsychological testing could have been done, giving the clinical conditions of patients. Patients were asked about history of anosmia during the acute phase. There were no clinical measurements of olfaction, due to the poor compliance of patients. Patients underwent FIM evaluation with a physiotherapist.[5] Some of these patients were subjected to a cerebral MRI with contrast or CT scan during their recovery, on the basis of clinical needs. In this case, three different investigators individually evaluated the cerebral images to assess cerebral trophism. Global cerebral atrophy (GCA) was evaluated on CT or MRI scans, based on fluid attenuated inversion recovery (FLAIR) acquisitions. Inter-observer results were standardized to obtain a single value.[19] GCA resulted from a systematic evaluation of 13 different regions of the brain to determine sulcal dilatation and dilatation of the ventricular system. At each level, a score of 0 (absent), 1 (mild), 2 (moderate) and 3 (severe) was given. The GCA resulted from the sum of all items (from 0 to 39).[19,20] White matter lesions (WML) were also analyzed on FLAIR acquisitions, with the Fazekas scale to assess white matter hyperintense lesions related to small vessels disease.[21] This scale consisted of a score of 0 (no or single WML), 1 (multiple WML), 2 (beginning confluence of WML), or 3 (large confluent WML).[21] A Fazekas score of 1 is considered normal in the elderly, while Fazekas 2 and 3 are usually considered pathologic. Scores of 2 or 3, when seen in normally functioning individuals, can be correlated with a high risk of disability.[22]

### Follow-ups

Patients were discharged and returned their homes after rehabilitation only with two consecutive negative swabs, at 24 hours interval. Patients were proposed follow-up (FU) visits, one month after hospital discharge. At FU, the following neuropsychological tests were performed: MMSE, MoCA, HRSD and the Davidson Trauma Scale[23] (DTS) to investigate possible post-traumatic stress syndrome.

### Data analysis

Patients were divided in four groups according to the respiratory support they received in the acute phase of the disease. Oxygen saturation (SpO2), partial pressure of carbon dioxide (PaCO2), pH and respiratory rate were used to define the respiratory intervention. Group 1 included patients who benefited from orotracheal intubation and ventilation from one to twenty-seven days (mean duration of intubation 12.39 ± 6.51 days). The following criteria were used to decide intubation and ventilation: tachypnoea, (RR>35), tachycardia, fatigue, agitation, use of accessory muscles, intercostal recession, SpO2>90, PacO2 > 60 mmHg, pH < 7.30. Group 2 included patients who benefited from non-invasive ventilation (NIV) using Biphasic Positive Airway Pressure (BiPAP or CPAP). Group 3 included patients who received oxygen therapy with Venturi Masks or reservoir Masks. Oxygen therapy was started with the aim to maintain SpO2 >90%, first, with Venturi Masks up to fiO2 50% (fraction of inspired oxygen), then with reservoir Masks up to fiO2 70%. Group 4 did not receive any oxygen therapy during the acute phase of the disease.

### Statistical analyses

Data from the different groups were analyzed using either one-way ANOVA or Kruskal-Wallis test, depending on the normality of data distribution, as evaluated by the Shapiro-Wilk test. Post-hoc analyses were performed using t-tests for independent values or Mann-Whitney analyses, depending on normality of the data. Bonferroni correction for multiple testing was applied. Spearman non-parametric correlation tests were used to investigate correlations between two variables. Follow-up data were compared to data at admission using Wilcoxon analyses. Data were considered significant when p<0.05. The commercially available software IBM SPSS Statistics v.23 (IBM Corp. ©) was used for all statistical tests.

## RESULTS

### Patients’ description

About 140 patients have been admitted in the COVID-19 Rehabilitation Unit during the above-mentioned period (about 80 days). These patients represented about 20% of the total number of COVID-19 patients hospitalized in the acute care units of the San Raffaele Scientific Institute (Milan, Italy). Out of these 140 patients, 87 met the inclusion criteria and were included in the study. Out of the 87 patients (62 Male, 25 Female, mean age 67.23 ± 12.89 years), 31 belonged to Group 1 (five Female, 26 Male, mean age 59.90 ± 8.92 years), 18 belonged to Group 2 (four Female, 14 Male, mean age 72.61 ± 8.15 years), 29 were included in Group 3 (14 Female, 15 Male, mean age 73.17 ± 12.19 years) and nine were included in Group 4 (two Female, seven Male, mean age 62.56 ± 20.06 years). Kruskal-Walis analyses indicated a significant effect of the main factor age (p<0.001). Indeed, the mean age of Group 1 was significantly lower than the mean age of Group 2 (p<0.006) and Group 3 (p<0.006).

### Neuropsychological data – infectious patients

The analyses of the MoCA scores showed that 74.2% of patients from Group 1, 94.4% of patients from Group 2, 89.6% of patients from Group 3 and 77.8% of patients from Group 4 presented with deficits, as shown by the analysis of the total score. One-way ANOVA on the main factor “total score” showed significant group differences (p=0.006) (Table 1). Specifically, Group 1 presented with higher scores compared to Group 3 (p=0.005, see Figure 1)). Significant differences between these two groups were observed in the sub-domains of short-term memory (p=0.010), attention (p= 0.016), abstraction (p=0.024), long-term memory (p=0.005), space and time orientation (p=0.034).

**Table 1:**
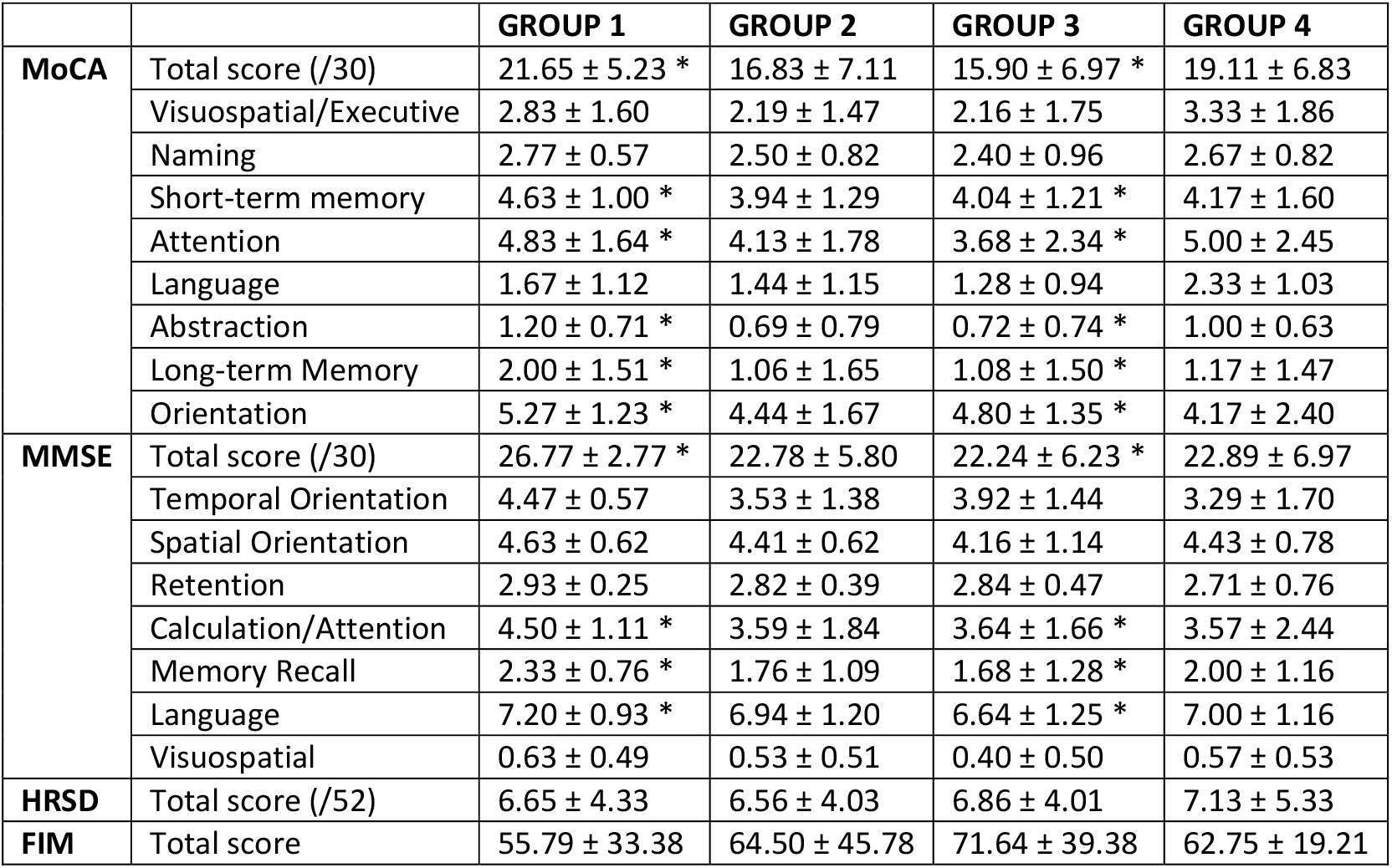
Neuropsychological and FIM evaluation of COVID-19 patients. Scores are divided according to each Group. * represents significant differences between groups (p<0.05). MoCA = Montreal Cognitive Assessment. MMSE = Mini Mental State Examination. HRSD = Hamilton Rating Scale for Depression. FIM = Functional Independence Measure.

**Figure 1:**
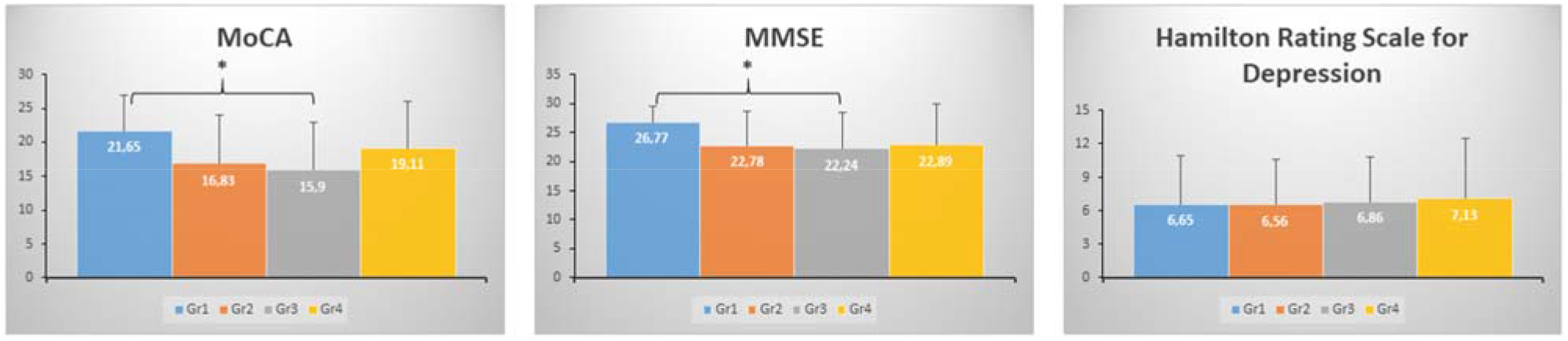
MoCA, MMSE and HRSD total scores. Figure 1 represents the values of the MoCA, MMSE, and HRSD total scores according to the different groups. * = p<0.05. HRSD= Hamilton Rating Scale for Depression.

MMSE analyses showed that 12.9% of patients from Group 1 had mild to severe deficits; 55.6% of patients from Group 2 had mild to moderate deficits; 48.3% of patients from Group 3 had mild to severe deficits; and 44.4% from Group 4 presented with moderate deficits. Kruskal-Wallis analyses showed significant differences between groups (p=0.021, Figure 1). Group 1 presented with higher scores compared to Group 3 (p=0.024). These differences were significant in the attention and calculation domain (p=0.003), in the memory domain (p=0.017), and in the language domain (p=0.024). There was also a trend towards differences in the temporal orientation sub-domain (p=0.051). Taken all groups together, MMSE total scores and MoCA total scores correlated significantly with patients’ age (MMSE: R=-0.262, p=0.014; MoCA: R= −0.376, p<0.001, see Figure 2).

**Figure 2:**
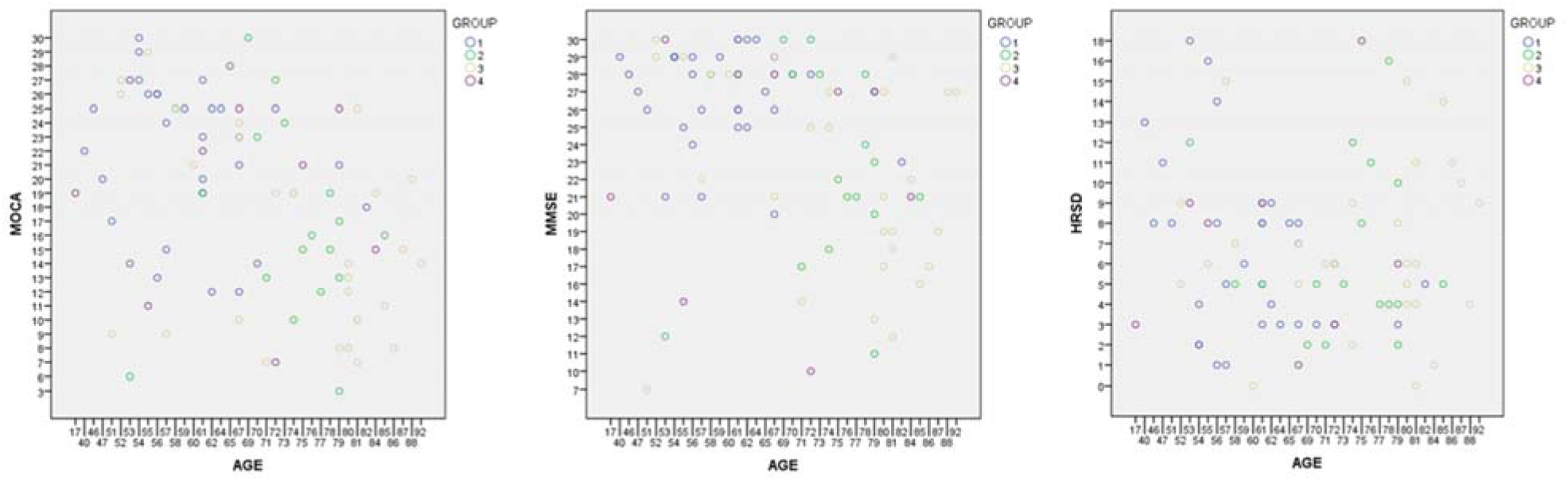
Neuropsychological values according in the four different groups according to patients’ age. Figure 2 represents the dispersion of MoCA, MMSE, and HRSD total scores among the different groups according to patients’ age. MoCA = Montreal Cognitive Assessment. MMSE = Mini Mental State Examination. HRSD = Hamilton Rating Scale for Depression.

Hamilton Rating Scale for Depression showed that 45.2% of patients from Group 1 had mild to moderate depression, 33.3% of patients from Group 2 had mild depression, 37.9% of patients from Group 3 had mild depression and 44.4% of patients from Group 4 had mild to moderate depression. There were no significant differences between groups in terms of total scores (p>0.05, see Table 1). There was no significant correlation between patients’ age and depression score (R= −0.048, p=0.663, see Figure 2).

### One-month follow-up

56 patients (22 of Group 1, 12 of Group 2, 20 of Group 3 and 2 of Group 4) underwent the follow-up evaluations, one month after discharge.

MoCA analyses showed that, at FU, 12 out of the 22 patients from Group 1 (54.5 %) had deficits; 10 out of the 12 patients from Group 2 (83.3%) had deficits; 17 out of 20 patients of Group 3 (85%) had deficits; and the 2 patients from Group 4 presented with deficits (100%). Although many patients still presented deficits at the MoCA evaluation, MoCA total scores at FU were however significantly higher than at admission (p=0.009). Conversely, MMSE analyses showed that, at FU, only 2 out of 22 patients from Group 1 (9.1%) had mild deficits; 1 out of 12 patients from Group 2 (8.3%) had mild deficits; 7 out of 20 patients of Group 3 (35%) had mild to moderate deficits; and 1 out of the 2 patients from Group 4 presented with moderate deficits. In concordance with such clinical improvements, MMSE total scores at admission and at 1-month FU differed significantly (p=0.004).

Regarding the analyses of HRSD at FU, 4 out the 22 patients of Group 1 (18.2%) had mild depression, 4 out the 12 patients of Group 2 (33.3%) had mild to moderate depression, 9 out the 20 patients of Group 3 (45%) had mild depression and none of the 2 patients of Group 4 had depression. Depression scores did not differ between FU and time of admission (p=0.167).

Lastly, at FU, 24 patients reported post-traumatic stress disorder symptoms at the Davidson Trauma Scale (12 patients from Group 1, 4 patients from Group 4, 7 patients from Group 3 and 1 patient from Group 4), while 32 reported no disturbances at all.

### Neurological signs

Eighteen out of the 87 patients had presented signs of anosmia since the onset of the disease. There were no significant differences in cognitive functions between the group of patients with anosmia and the group of patients who did not suffer from anosmia, either for the MMSE or the MoCA total score (p=0.555 and p=0.410, respectively). FIM scores did not differ either between anosmic and non-anosmic patients (p=0.771). FIM scores did not significantly differ between groups (p>0.05, see Table 1), nor correlated with age (R= −0.048, p=0.187).

Thirty-seven patients underwent CT head scans or cerebral MRI (14 patients in Group 1, five patients in Group 2, 12 patients in Group3 and six patients in Group4). Total CGA scores ranged from 0 to 25 (mean score 8.16 ± 5.85). Patients from Group 1 had a mean CGA score of 5.29 ± 3.50, patients from Group 2 had a mean score of 9.6 ± 3.78, patients from Group 3 had a mean score of 11.42 ± 7.59, and patients from Group 4 a mean score of 7.17 ± 4.96). Kruskal-Wallis analysis showed that there were no significant differences between the four groups (p=0.09). Taken all patients together, there were significant correlations between total CGA scores and the factors age (R= 0.582, p<0.001) and MoCA total score (R=-0.339, p=0.040). There were no significant differences in GCA scores between patients with or without anosmia (p=0.805).

## DISCUSSION

In this communication, we analyzed the neuropsychological data of a group of 87 infectious COVID-19 patients, in the sub-acute phase of the disease, out of about one thousand patients admitted to the ER of our Institution in the last four months. These patients presented with clinical indications for functional rehabilitation (FIM<100) shortly after the onset of the disease and immediately after the acute phase of the infection. The great majority of patients were men (71 %). All four groups of patients presented similar degrees of depression, with about 40% of patients reporting mild to moderate depression. Moreover, more than 80% of these patients presented with cognitive deficits, as shown by the MoCA analyses. Differential deficits were observed between groups. Indeed, most of these COVID-19 patients had previously benefited from oxygen therapy (57 out of 67), with various degrees of invasiveness. Among patients who previously received oxygen therapy, Group 1 benefited from invasive ventilation and sedation. This group presented with better cognitive status compared to the other groups. There were especially significant differences between this group of patients and Group 3, who received oxygen therapy through Venturi Masks. The cognitive impairments resulted mainly in deficits in short- and long-term memory, executive functions, abstraction, language, and orientation. These data indicate that patients who benefited from the most aggressive respiratory assistance had better preserved cognitive functions in the subacute phase of the disease. However, patients who benefited from the most invasive respiratory assistance were also the youngest. Indeed, Group 1 had a younger mean age compared to groups 2 and 3. This age-difference might have explained, at least in part, the differences in cognitive performance between Groups 1 and 3 (although no significant differences in cognitive functions were observed between Group 1 and Group 2). Cognitive data (MMSE and MoCA total scores) significantly correlated with age. These data indicate that the youngest patients were the ones who benefited from the most invasive respiratory assistance and the ones who showed the most conserved cognitive functions.

Anosmia was reported only in a minority of our patients (18 out of 87). Thus, this neurological sign does not appear to be relevant enough to be used as a screening tool for neurological involvement. Similarly, FIM scores did not differ between groups, showing that the entity of the cognitive deficit was not associated with motor deficits. This result shows that FIM evaluation should be associated with other neurological and neuropsychological testing to better identify neurological dysfunctions in COVID-19 patients. Similar cognitive deficits are often observed following acute respiratory distress syndrome (ARDS). [24] 70% to 100% of ARDS survivors would experience cognitive impairments at hospital discharge. [25,26] Low PaO2 has been associated with long-term cognitive impairment, especially in the domains of executive functions and psychomotor tasks. [27,28]

MoCA evaluation was more sensitive compared to MMSE to detect these cognitive impairments. As demonstrated in studies on the effects of ARDS, MMSE would have a poor sensitivity in detecting cognitive impairment after ARDS. [29] Thus, implementation of sensitive cognitive testing tools like MoCA may help better assess patients’ cognitive functions and, as a consequence, provide better care and functional recovery outcome. Our study showed that MoCA was sensitive enough to detect cognitive impairments of various domains in infectious COVID-19 patients. Other tools previously validated in ARDS studies included the Confusion Assessment Method for Intensive Care Unit patients (CAM-ICU), with a good sensitivity and specificity in detecting delirium [30], or the Modified Blessed Dementia Rating Scale and the Informant Questionnaire on Cognitive Decline in the Elderly to evaluate for pre-existing cognitive deficits. [31] None of our patients presented with delirium, as often observed in ARDS or in post-intensive care syndrome (PICS). [32–35] PICS can also be accompanied by long-term cognitive impairments, especially regarding memory, executive functions, language, attention, and visual-spatial abilities. [32,35] Thus, part of our results could be related to PICS, although we showed that patients who benefited from the most invasive respiratory assistance had better preserved cognitive functions and did not show worse mood alterations, compared to the other groups. No signs of psychosis, delirium, or other altered mental status have been observed in these patients.

Our neuropsychological data were unexpected as they showed that patients who underwent sedation and ventilation, i.e. patients in the most critical clinical state, were patients with the less compromised cognitive status. These results might be due differences in oxygen volume received during hospitalization. One might also hypothesize that sedation might have spared patients from the stress that such a critical illness might have induced in otherwise conscious patients. Indeed, acute and chronic stress would be associated with increased mechanisms of inflammation and enhanced attentional processing of negative information. Both phenomenon are predictive of depression symptoms that, in turn, increase inflammatory and cognitive stress reactivity. [36] At one-month follow-up, patients had partly recovered their cognitive impairments, especially as measured by the MMSE. MoCA scores were still showing deficits in most of the patients. These differences observed between MMSE and MoCA scores might be related to the higher sensibility of MoCA to detect slight variations in cognitive functioning, as shown by several studies on Alzheimer or Parkinson’s patients. [17,37] Moreover, 43% of patients tested at one-month follow-up had symptoms of PTSD, especially patients who underwent the most invasive treatments (orotracheal intubation). It had been shown that about 25% of patients admitted into ICU develop PTSD afterwards. [38] Such risk of developing PTSD has been recently shown to be higher in COVID-19 ICU survivors. [39] Thus, taken altogether, our results show possible cognitive impairments in the sub-acute phase of COVID-19 and that can persist even one month after discharge, demonstrating the need for psychological treatment and assistance of post COVID-19 patients.

## Conclusion

Our study focused on the sub-acute phase of the COVID-19 in positive patients with functional disability, in order to investigate the essential needs of clinical assistance for these patients. Like many healthcare centers worldwide, our Institute has been shaken by the dramatic Sars-CoV-2 pandemic wave. In the exponential phase of the pandemic in Italy (March/April 2020), about 60 patients were admitted in the ER daily. Therefore, all patients in need for respiratory assistance could hardly be admitted to the ICUs, and choices regarding the degree of invasiveness of assistance had to be done. Nowadays, at distance from the emergency pic, the data reported in this paper should serve as a demonstration of how the choice of respiratory assistance can influence cognitive functions of COVID-19 patients, especially in aging patients. Our data showed that about 80% of our patients presented with cognitive deficits in this sub-acute phase of the disease, about 40% of patients suffered from mild to moderate depression, and that these deficits were more important in the older patients. The decisions on respiratory assistance did not have differential consequences on the motor symptoms. Thus, these data showed that, in the acute phase of the disease, the most aggressive strategies for respiratory assistance did not have negative consequences on neuropsychological or motor outcome. Conversely, the age of patient appeared as a risk factor for neuropsychological impairment of COVID-19 patients. Since a high number of patients still had cognitive impairments at one month FU, our data also demonstrate the need for long-term psychological support and treatment for post COVID-19 patients.

## Data Availability

All relevant data are available in the manuscript.

